# Fusing Data from CT Deep Learning, CT Radiomics and Peripheral Blood Immune profiles to Diagnose Lung Cancer in a Cohort of Patients Experiencing Symptoms

**DOI:** 10.1101/2025.01.10.24319412

**Authors:** Rami Mustapha, Balaji Ganeshan, Sam Ellis, Luigi Dolcetti, Mukunthan Tharmakulasingam, Karen DeSouza, Xiaolan Jiang, Courtney Savage, Sheena Lim, Emily Chan, Andrew Thornton, Luke Hoy, Raymond Endozo, Rob Shortman, Darren Walls, Shih-hsin Chen, Mark Rowley, Anthony CC Coolen, Ashley M Groves, Julia A Schnabel, Thida Win, Paul R Barber, Tony Ng

## Abstract

**Background:** Lung cancer is the leading cause of cancer-related deaths. Diagnosis at late stages is common due to the largely non-specific nature of presenting symptoms contributing to high mortality. There is a lack of specific, minimally invasive low-cost tests to screen patients ahead of the diagnostic biopsy.

**Methods:** 344 patients experiencing symptoms from the lung clinic of Lister hospital suspected of lung cancer were recruited. Predictive covariates were successfully generated on 170 patients from Computed Tomography (CT) scans using CT Texture Analysis (CTTA) and Deep Learning Autoencoders (DLA) as well as from peripheral blood data for immunity using high depth flow-cytometry and for exosome protein components. Predictive signatures were formed by combining covariates using Bayesian regression on a randomly chosen 128-patient training set and validated on a 42-patient held-out set. Final signatures were generated by fusing the data sources at different levels.

**Findings:** Immune, CTTA and DLA single modality signatures had overall AUCs of 0.69, 0.70 and 0.73 respectively. The final combined signature had a ROC AUC of 0.81. The overall sensitivity and specificity were 0.72 and 0.77 respectively.

**Interpretation:** Combining immune monitoring with CT scan data is an effective approach to improving sensitivity and specificity of Lung cancer screening even in patients experiencing symptoms.

**Funding:** CRUK [C1519/A27375], Wellcome Trust/EPSRC Centre for Medical Engineering [WT203148/Z/16/Z], NIHR Clinical Research Facility at Guy’s and St Thomas’ NHS Foundation Trust, NIHR Biomedical Research Centre.

**Research in Context:** *Evidence before this study:* Lung cancer is the leading cause of cancer related deaths and previous studies have shown that early diagnosis is often difficult and one third of patients return to their GP three or more times before a referral to a specialist according to a study in the British Journal of Cancer in 2013. Screening is a possible solution and low dose CT scans for patients with high-risk has been implemented, but this excludes non-smokers and the young and can be inaccurate according to studies in 2022 and 2023. In addition, the workload of screening on radiologists is resulting in delays in examinations as recently reported by the Royal College of Radiologists in 2024. Some studies have suggested the benefit of a blood test, to augment a CT scan, based on DNA fragments or circulating tumour cells, for example (2021-2023). A PubMed publications database search (search term: “blood“[Title] AND “CT“[Title] AND “Lung“[Title] AND “cancer“[Title]) revealed 17 results on 8^th^ May 2025, of which 2 were directly relevant involving combining a PET/CT scan with inflammatory blood measurements, rejecting those where blood referred to blood volume or perfusion imaging or ctDNA.

*Added value of this study:* We propose a blood test based on the immune response to the presence of cancer (in contrast to other studies which are based on detecting nucleic acids) that is combined with the automated analysis of CT scans (reducing radiologist workload) by machine learning, which may provide a route to screening patients with symptoms of lung diseases. The advantage of a blood test based on the immune response is that immune cell-based detection of cancers can occur <u>when the tumours are</u> <u>relatively small and at an earlier time point</u> than the shedding of nucleic acids. We recruited 344 patients between October 2020 and November 2021 into the LungExoDETECT study, with a blood test for immune profiling and routine CT scan. After following their progress to determine their cancer diagnosis, we were able to make a mathematical model that predicts diagnosis from 12 measurements based on the blood and CT scan, and which was validated in a patient subset.

*Implications of all the available evidence:* Our study has important implications for the field: (1) We reinforce previous our previous evidence that the response of the immune system to cancer can add additional information for detecting cancer to that provided by the automated analysis of CT scans. (2) Advanced Bayesian machine learning can produce simple mathematical models that predict the presence of cancer and are highly interpretable in terms of why a particular prediction has been made by the model by pinpointing the particular component of the immune system that contributes to the prediction.

## Introduction

### Lung cancer and importance of early detection

Lung cancer is by far the leading cause of cancer-related deaths worldwide with an estimated 1.8 million death in 2020 ^1^. In the U.S.A., lung cancer accounts for about two and a half times the number of cancer deaths than the second leading cause, colorectal cancer. This is despite the fact that lung cancer only contributes to about 12% of newly diagnosed cancer cases, which is far less than prostate cancer and breast cancer, the leading cancer diagnoses for men and women, respectively ^2^. This high mortality of lung cancer has been linked to diagnosis at late stages. 48% of lung cancer cases are diagnosed with a distant metastasis and an expected 5-year survival rate of 8% compared to the ≥60% rate when diagnosis happens at a localized stage ^3^. In England, 5-year survival for a stage I diagnosis is 63%, which drops down to 4.3% for a stage IV diagnosis ^4^. Delayed diagnosis can be attributed to the largely asymptomatic nature of early-stage lung cancer ^5^. Other data have shown that patients delaying their first general practitioner (GP) visit after manifestation of early symptoms is another obstacle to a timely diagnosis ^6^. Beyond that, a study from 2013 showed that a third of patients with lung cancer have 3 or more visits with their GP with lung cancer associated symptoms before special referral, which forms a sharp contrast to the 3% observed in patients with breast cancer^7^. Similarly, the UK’s National Cancer Diagnosis Audit reported primary care delays of 60 and 90 days experienced by 17.9% and 10.8% of patients with suspected lung cancer, respectively ^8^. This highlights the need for lung cancer screening, which to a certain extent has been implemented in the form of low dose computed tomography (LDCT) for patients of high-risk. Despite this practice leading to about a 20% reduction in mortality across multiple clinical trials, there is still room for improvement ^9, 10^. LDCT screening excludes patients that are younger and those at lower risk as it has been shown to be not reliable as a screening tool for the broader population, owing to its high chance of false positives ^11, 12^. Lung cancer screening also heavily relied on smoking history, which excludes the early detection of lung cancer in non-smokers which represent around 25% of cases ^13^. There is a clear need for a minimally invasive robust screening test for lung cancer with similar sensitivity and specificity to a mammography for breast cancer, or the faecal occult blood test currently in use for colorectal cancer. Furthermore, this needs to be extended beyond screening, as numerous healthcare systems, including the NHS in the UK, have put in place a rapid diagnostic pathway for patients already symptomatic and suspected of lung cancer ^14^.

### Role of radiologist and importance of alternative image analysis

The diagnostic pathway for lung cancer is heavily reliant on radiologist assessment at multiple stages. According to the UK NHS National Optimal Lung Cancer Pathway (NOLCP) ^15^ and National Institute for Health and Care Excellence (NICE) guidelines, the typical pathway may include a chest radiograph (CXR), a CT scan, a PET-CT, a CT guided biopsy, all of which require a radiologists intervention, bronchoscopy (EBUS-TBNA, EUS-FNA or navigational) and pleural procedures. This reliance is further exacerbated by the large amount of CT imaging data from lung cancer screening programmes. This workload combined with the shortage of radiologists in the UK and other European countries is manifesting in delays in examinations as recently reported by the Royal College of Radiologists ^16, 17^. Implementing automated image analysis approaches like radiomics-based texture and deep learning analyses is a viable approach, especially given the availability of large screening datasets for training purposes. The low-dose radiation used in LDCT, to minimize patient exposure, does negatively impact the visibility of small or low-contrast lesions due to increased noise and reduced spatial resolution and artificial intelligence can alleviate this in many areas of CT screening ^18,19^. In this study we exploit the latent vector of a deep autoencoder as a low dimensional representation of the lesion image characteristics and evaluate its predictive performance.

### Liquid biopsy

Using a liquid biopsy for early detection and diagnosis of lung cancer has been heavily assayed to varying degrees of success ^20^. Of note, Galleri® which is a circulating tumoral DNA (ctDNA) methylation-based multicancer early detection (MCED) test is seeing adoption across clinics. Early results seem very promising in the symptomatic setting, though these are not comparable across all cancer types. Of note, sensitivity for stage I lung cancer was 8.7% and 0% in the symptomatic and asymptomatic settings, respectively ^21, 22^. Combining LDCT with minimally invasive blood test is one approach that can improve the shortcoming of either approach. This has proven useful in the BioMilk study that showed that double positivity LDCT and microRNA signature doubles the sensitivity of LDCT alone ^23^. Similarly, other studies combined circulating proteins with LDCT achieving improved specificity ^24^. However, there is still room for improvement using other blood biomarkers which include circulating tumour cells, microRNA, exosomes, tumour educated platelets, metabolites, tumour associated proteins, and autoantibodies. In addition, peripheral cell-based flow cytometry immune markers ^25–27^ (especially the immune checkpoint receptors) can potentially provide treatment targets for the cancers detected.

### Immunology in lung cancer

Little research exists on the feasibility of immune profiling of circulating immune cells as a biomarker for early lung cancer. Peripheral blood mononuclear cells (PBMCs) are a heterogenous population of immune cells usually classified into subsets. The numbers, relative frequencies and functional status these subsets provide information on the state of the immune response and there have been recent advents in technology beyond classical multiparametric flow cytometry. Using patient PBMCs, single cell RNA sequencing was used to identify circulating anti-tumour CD8 cells, and Cytometry by time of flight (CyTOF) was used to correlate the frequency of HLADR expressing monocytes with response to ICB therapy ^28, 29^. Numerous research groups, including ours, have shown that changes at the tissue level of a solid cancer immune response are reflected as systemic changes in the immune response that can be detected at the peripheral level ^28–30^. Lung cancer promotion after initial mutagenesis has been shown to be inflammation driven, with a recent link established to air pollution ^31^, further highlighting the potential of an immune based biomarker for early detection. Cancer specific T cell receptor repertoire has been shown to be effective at identifying early-stage lung cancer in an LDCT screened cohort. This proves that cancer specific immune cells can be reflected in the periphery ^32^.

## Methods

### Study design and data collection

The study entitled “Improving the Early Detection of Lung Cancer by Combining Exosomal Analysis of Hypoxia with Standard of Care Imaging (LungExoDETECT)” (https://clinicaltrials.gov/ct2/show/NCT04629079) is a prospective cohort study of patients referred to secondary care for the investigation of clinical symptoms or signs suspicious of lung cancer. The study analysis will determine whether the assay can detect clinical lung cancer at the time of imaging, and interval cancers during subsequent follow up. The study aimed to establish preliminary ROC AUC and sensitivity/specificity data for the “combined CT/blood risk stratification marker” and provide initial data on the potential association of the “combined CT/blood risk score” with the subsequent cancer progression and treatment response. The gold standard lung cancer diagnosis was determined by the National Optimal Lung Cancer Pathway (NOLCP).

The study included patients who have been referred to the Lung Cancer Clinic and Multi-Disciplinary Team (MDT) at The Lister, Hertford County, UK and New QEII Hospitals, Welwyn Garden City, UK for investigation of suspected lung cancer. Participants were recruited between October 2020 and November 2021 (344 patients). As part of the standard of care pathway they received a CT scan, at that time peripheral blood samples were collected. 174 cases were excluded, 162 due to lack of visible lesion or lesion <6mm and 12 cases with non-lung cancer. Sex/gender was self-reported by study participants and is reported but not included in multivariate analysis. The patient pathway and study design is illustrated in Figure 1 and patient characteristics are in Table 1.

**Figure 1.**
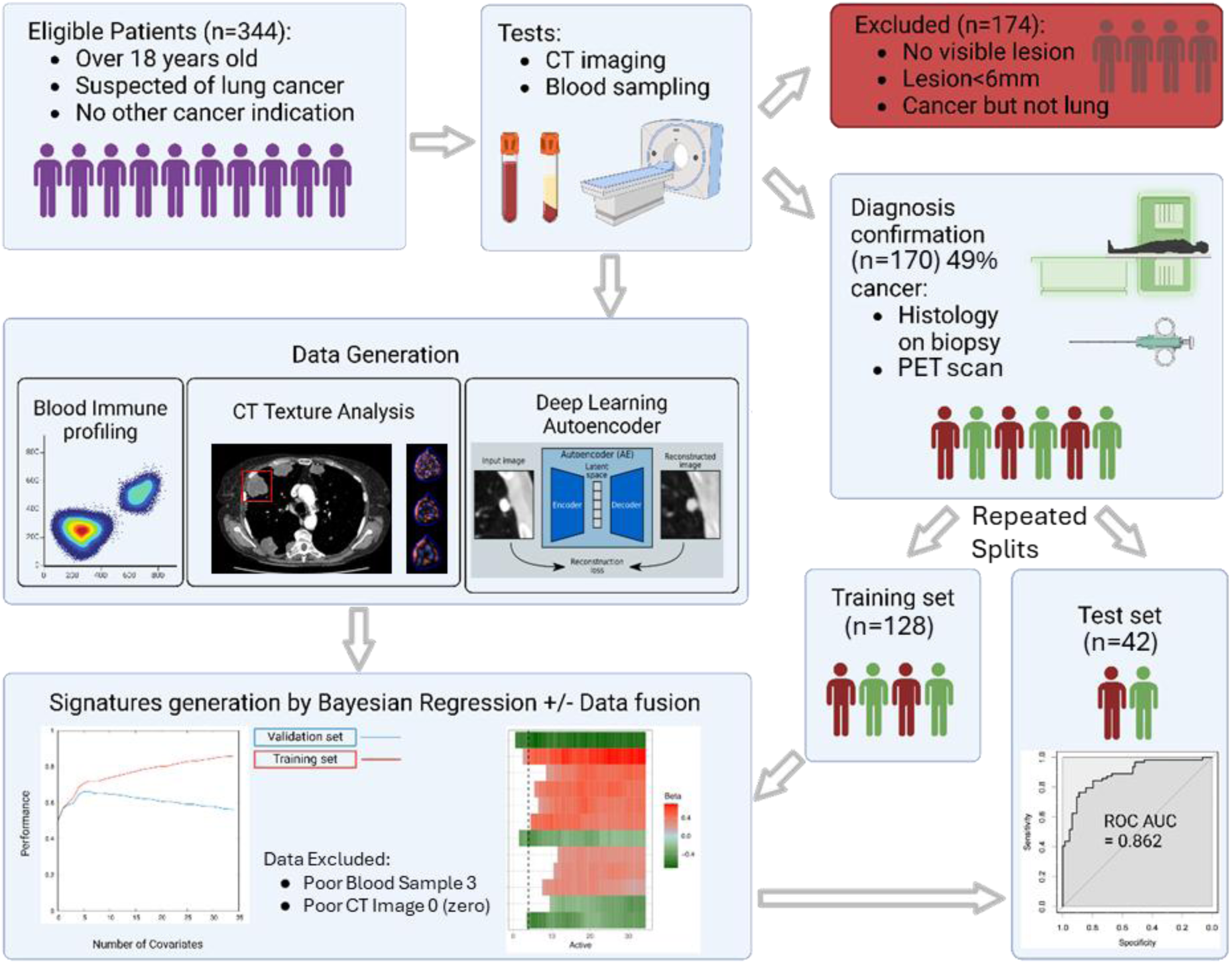
Showing the patient pathway and study design. 344 patients referred to the clinic were recruited to the study. As part the standard of care pathway they received a CT scan, at which time we collected the peripheral blood samples. 174 cases were excluded from the analysis set due to lack of visible lesion, the lesion was smaller than 6mm or the diagnosis was a non-lung cancer diagnosis. The analysis was performed on the remaining patients.

**Table 1.** Patient characteristics and distribution. The table shows the difference between cancer and non-cancer as well as staging for patients with cancer. *P-values were calculated to compare data from individuals with and without lung cancer for the following variables: mean ages using Student’s unpaired two-tailed t-test whereas for sex distribution, smoking status and cohort using a χ2 test.

| Patient Characteristic | Non-cancer<br>*n* = 91 | Lung cancer<br>*n* = 79 | p-value* |
| --- | --- | --- | --- |
| Age |  |  |  |
| Mean | 71.9 | 74.9 | 0.152 |
| Range | 27.0 to 94.0 | 33.0 to 97.0 |  |
| Sex |  |  |  |
| Male | 43 | 38 | 0.646 |
| Female | 48 | 41 |  |
| Smoking status |  |  |  |
| Ex-Smoker | 33 | 31 | 0.07 |
| Non-Smoker | 29 | 12 |  |
| Smoker | 17 | 21 |  |
| Unknown | 12 | 15 |  |
| Cohort |  |  |  |
| Training | 64 | 64 | 0.1 |
| Test | 27 | 15 |  |
| Staging |  |  |  |
| No Cancer | 91 |  |  |
| Stage I |  | 23 |  |
| Stage II |  | 9 |  |
| Stage III |  | 18 |  |
| Stage IV |  | 29 |  |

### Sample and Data Processing

The details of CT scan acquisition, CT Texture Analysis (CTTA) (Supplementary Figure 1, Supplementary Figure 2), the Deep Learning Autoencoder (DLA), peripheral blood sampling and processing, flow cytometry and model (aka signature) generation are given in the Supplementary Methods. The total number of features generated by each modality are: 42 from CTTA, 32 from DLA, 45 from flow cytometry and 68 from exosome dot-blots (Supplementary Table S1, Supplementary Table S2, Supplementary Table S3).

### Model Building and Performance Analysis

Separately, using the data from each analysis, a risk signatures (models) were generated using Bayesian multi-variate regression (BMR) which also performed covariate selection (supplementary material). Although 170 is a reasonable number of samples it is not significantly larger than the combined number of covariates. It is therefore reasonable to use all 170 for model building and through bootstrapping training and test sets, gain indicators of performance. Recent literature indicates that these BMR approaches will not overfit and therefore will generalise under the assumption that the training set is representative^33, 34^.

Of all the study samples, one quarter was randomly held out of the model training and served as an internal test set. This was repeated 8 times to get a good estimate of future performance independent of the training/test splitting. The performance of both univariate biomarkers and model predicted outcomes were judged by the area under the receiver operator characteristic curve (ROC AUC) with the cancer diagnosis result from the clinic as the gold standard using the R ‘pROC’ package and the ‘ci.auc’ function. Sensitivity and specificity were calculated at the Youden point of the ROC. A Risk Score can be generated by combining covariate raw values in a linear model according to their given weights and intercept. Final signatures were generated from covariates that were significant in at least 50% of the 8 repeated training runs, and assed on the full cohort. This offers credence to the robustness of the selected features.

### Statistics

For comparing groups, a Student’s unpaired two-tailed t-test or a χ2 test was used for continuous or discrete variables, as these are well used and understood tests for this purpose. Hazard Ratios, P-values and confidence intervals from multivariate analyses were calculated from posterior parameter probability distributions. In the Bayesian framework these are calculated using exact expansion methods, that (whenever required by the data) go beyond quadratic (i.e. Fisher matrix based) approximations, and are easily interpretable in line with other tests.

### Ethics

All procedures were performed in compliance with relevant laws and institutional guidelines and have been approved by the appropriate institutional committee(s), and informed consent obtained from the subjects (REC reference: 19/EE/0357 20^th^ Feb 2020).

## Results

### Patient Characteristics

The characteristics of the patients suspected of lung cancer and included in the final analysis set are shown in Table 1. Of those suspected of having lung cancer 46.5% (79 patients) were diagnosed with lung cancer over the follow up period. There were no significant differences between the patients with and without cancer with regards to sex, age, smoking status nor assignment to training or test set. The division of patients between training (3/4) and test sets (1/4) was retrospective and at random, with the condition of having a balance of patients with and without cancer representative of the whole cohort (Supplementary material). Amongst the patients with lung cancer, the division across stage is shown in Table 1. Of note, 23 of the lung cancers (29.1%) were stage I, signifying that this is a good cohort for the potential development of a screening signature. 12 patients of the total recruited were diagnosed with a cancer that is not lung cancer (characteristics in Supplementary Table S4).

### Generation of a Predictive Model from CTTA and DLA

The imaging data were analysed using CTTA and DLA, as explained in the methodology. The signatures are presented in Figure 2A. The DLA signature performed better than the texture analysis signature on both the training and test cohorts. On the training sets DLA had a range of AUC of 0.72-0.77 (mean ± 1 standard deviation), whereas CTTA had an AUC range of 0.67 to 0.73. On the test sets DLA had an AUC range 0.56 to 0.71, whereas CTTA had an AUC range of 0.57-0.79 (Figure 2A). Figure 2A also represents the result of combining CTTA and DLA analyses.

**Figure 2.**
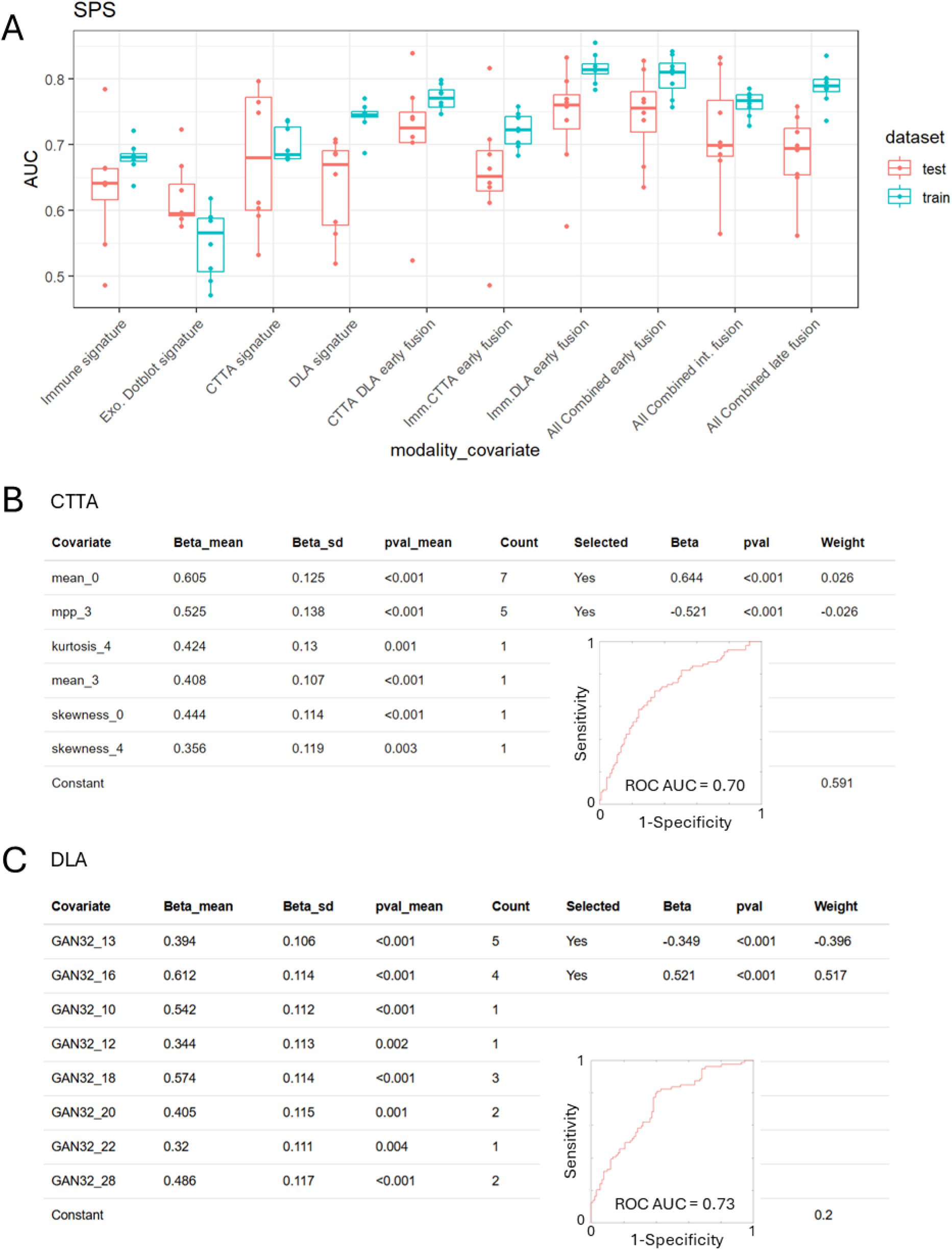
Risk generation by BMR from repeated data splits and comparing CT texture analysis to deep learning autoencoder. (A) Box plots showing the distributions (mean and inter-quartile range, points overlaid) of AUCs obtained from 8 random training/test splits of the data. (B, C) Outcome association statistics for CTTA and DLA of the 8 repeats, and the count of the number of times the covariate was indicated as important. Covariates indicated more than 50% of the time were selected for the final model generation; the final association (Beta), p-value for the association and weight in the final signature are given, as well as a ROC curve and AUC value for the final signature.

The CTTA features consistently selected by BMR across the repeated training splits were the mean intensity at spatial scale filter (SSF) = 0 (Mean_0) – which relates to the CT attenuation / density of the tumour, and the mean intensity of positive pixels at scale SSF=3 (mpp_3) – which relates to the average brightness at fine texture scale (corresponding to features of 3mm in size) within the tumour.

A preliminary exploration of the interpretation of the DLA latent space features selected by BMR was performed. This involved passing a lesion image into the encoder before manually varying the value of predictive features. By varying the value of a feature in this way, and passing the resulting latent space codes through the decoder, the impact of each feature in image space could be viewed. The 5 predictive DLA features selected by BMR (numbered 13 and 16) were observed to relate to lesion size, morphology (e.g. spiculation and sphericity), location within the lung (evidenced by changing distance and orientation of pleura), and degree of pleural attachment (Supplementary Figure 3). As quantitative evaluation of these interpretations would require large annotated datasets, including novel quantitative measures for characteristics such as pleural attachment, we leave this for future work.

### Generation of a Predictive Model from Peripheral Blood

The peripheral blood samples were analysed for their immune and their exosome protein content (dot-blot) as indicated in the methods. Separately, using the data from each analysis modality, a risk signature was generated (Figure 3A). The flow cytometry signature performed far better on the training and test cohort than the exosome protein content signature. On the training sets, the immune signature had an AUC range of 0.66-0.70, whereas the exosome derived signature had an AUC range of 0.50-0.60 (Figure 2A). Of note, the flow cytometry signature was driven by two significant features: a high proportion of type 2 dendritic cells indicated a non-cancer pathology, whereas a high proportion KIR3DL1 expressing CD8 T lymphocytes indicated lung cancer (Figure 3A). The exosome dot-blot results were not consistent, with the exception of some consistency of PD1, and was not used for further combined risk signature generation (Figure 3B).

**Figure 3.**
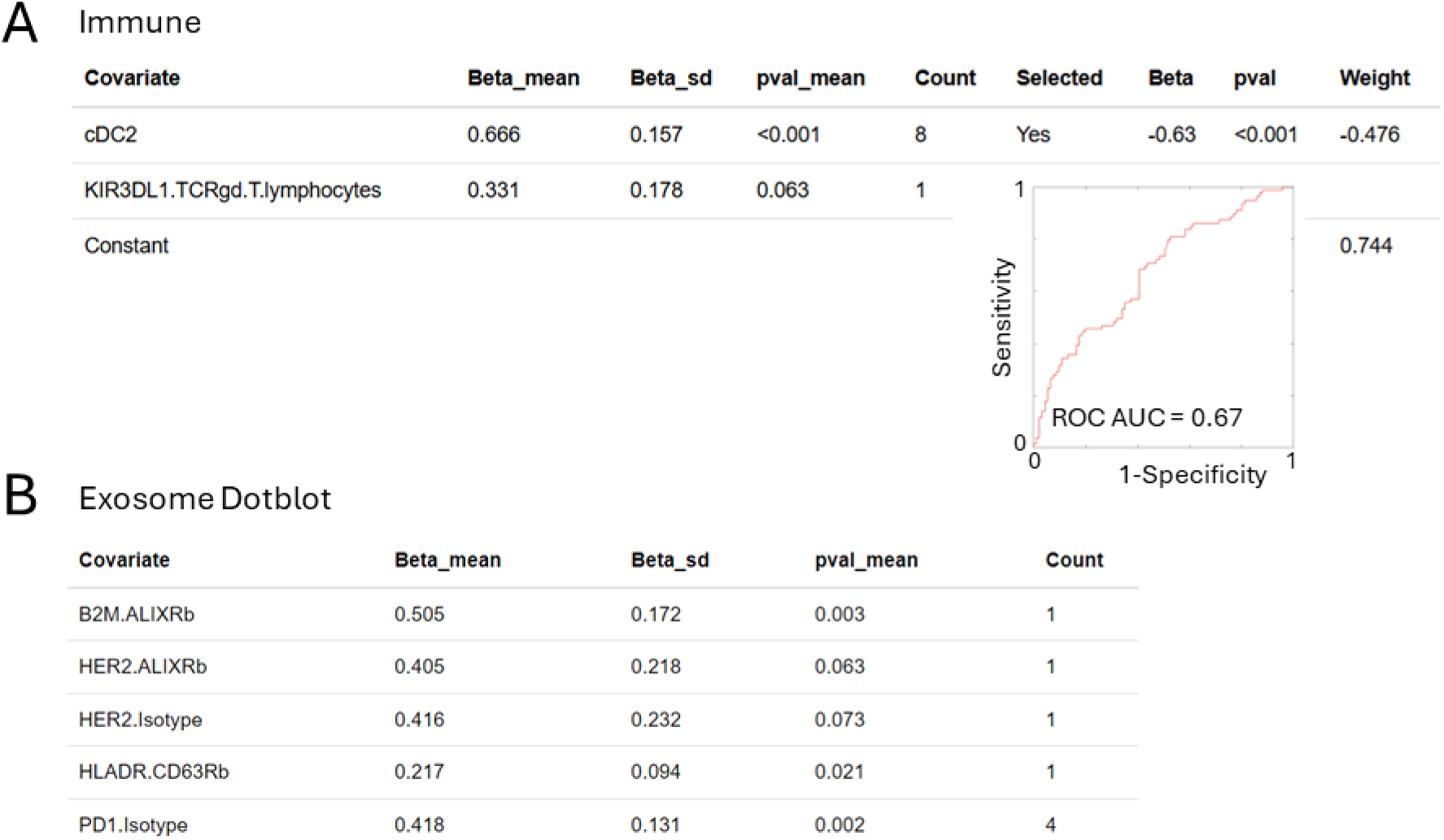
Risk generation by BMR from peripheral blood samples comparing flow cytometry to plasma exosome dotblot analysis. (A, B) Outcome association statistics for Immune and Dotblot of the 8 repeats, and the count of the number of times the covariate was indicated as important. Covariates indicated more than 50% of the time were selected for the final model generation; the final association (Beta), p-value for the association and weight in the final signature are given, as well as a ROC curve and AUC value for the final signature. Associations for the dotblot data were so low that a final model was not generated.

### Combining CT Analyses with Peripheral Blood Data

We proceeded to combine the data from the blood sample with the CTTA and DLA individually, at the covariate level (early fusion). Both combined signatures from BMR featured covariates from the immune and imaging data with the upregulation of KIR3DL1 CD8 T cells being the strongest predictor of a lung cancer (Figure 4A). The risk signature generated from the combination of flow cytometry and DLA performed better on the training set with an AUC range of 0.77-0.83, which transferred well to the test sets: 0.68-0.81 (Figure 2A). The risk signature generated from the combination of peripheral immunity and CTTA on the training set had an AUC range of 0.79-0.84, which also transferred well to the test sets: 0.66-0.82. Final models were generated from the consistent covariates with AUCs of 0.81 and 0.71 (Figure 4).

**Figure 4.**
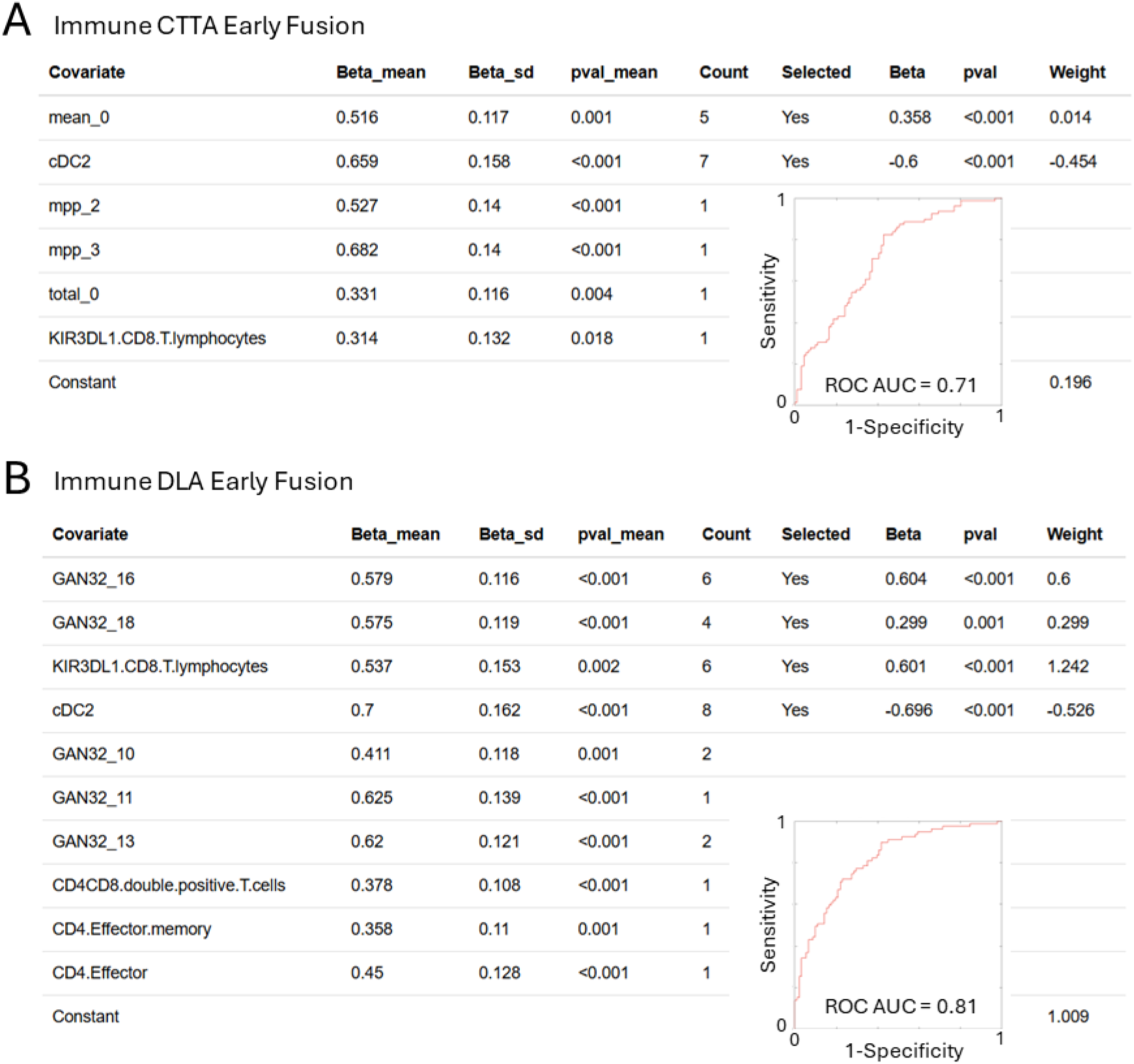
Risk generation by BMR from CT image analysis combined with flow cytometry, comparing deep learning and texture analysis. (A, B) Outcome association statistics for Immune-CTTA and Immune-DLA fusion models of the 8 repeats, and the count of the number of times the covariate was indicated as important. Covariates indicated more than 50% of the time were selected for the final model generation; the final association (Beta), p-value for the association and weight in the final signature are given, as well as a ROC curve and AUC value for the final signature.

### Generation of a Combined Model by Data Fusion

We proceeded to combine all methods of CT and blood analyses using data fusion at different levels. Combination at the covariate level (early fusion) did not show a major improvement of the risk signature with an AUC range 0.77-0.83. The signature generation process included co-variates from all three modalities, but no CTTA covariates were selected in 50% of runs (Figure 5A).

**Figure 5.**
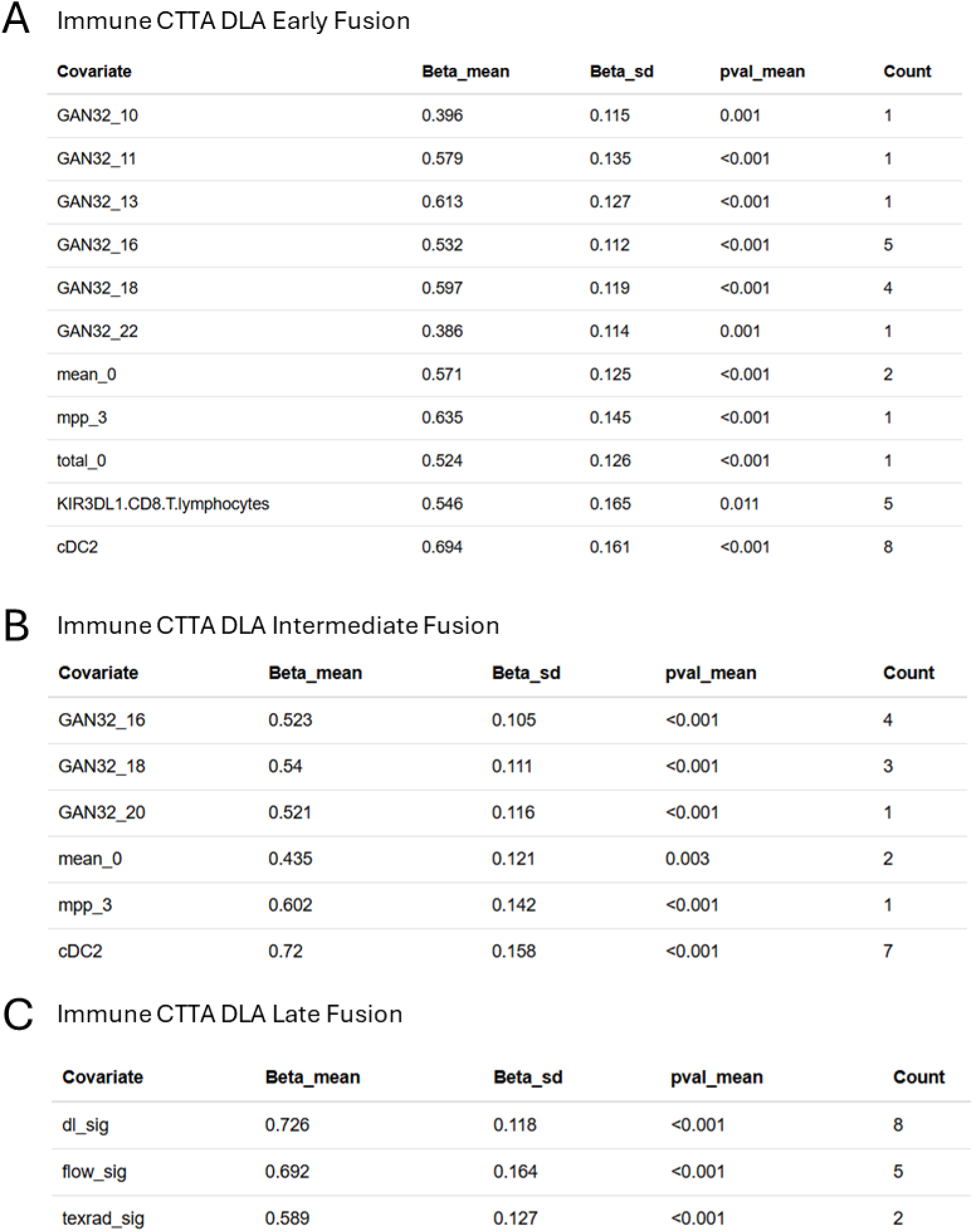
Risk signature generation from fusion models. (A, B, C) Outcome association statistics for Immune-CTTA-DLA early, intermediate and late fusion models of the 8 repeats, and the count of the number of times the covariate was indicated as important.

We also generated predictive signatures by fusing the features included in individual signatures (feature level or intermediate fusion) and by fusing the individual signatures themselves (signature level or late fusion, Figures 5B,5C). These did not show better performance than the early fusion models.

An equivalent approach to data combination and signature generation was performed using an Elastic Net algorithm (glmnet R package) ^35, 36^ which produced some better signatures on the training sets, but struggled with overfitting which impacted the ROC AUCs on the test set. We are focusing on the results from BMR since they are more consistent and offer less overfitting. A summary of all the Elastic net signatures is in Supplementary Figure 4. Overcoming the problem of overfitting was the main reason for the use of the Bayesian regression, which will optimally choose the number of covariates to be predictive given the evidence in the training set. This is in contrast to the commonly used Elastic Net algorithm, which in its LASSO form will select an optimal set of parameters through regularisation but not information content.

### Combined Signature Specificity and Sensitivity to Detect Lung Cancer at Different Stages and Across Other Cancers

Disease staging was intentionally excluded from the data sets for both signature generation and testing. As current methodologies struggle at detecting early-stage lung cancer ^21, 22^ where current available therapies are most effective, we assessed the ability of the final signature to detect cancer across all stages for the training and test sets (Figure 6A). The overall sensitivity was 72%. The sensitivity was high in most Stages at 76-78%, with a drop in Stage II but we have very limited numbers in that Stage. Interestingly, the specificity was high at 77% despite the fact that these are patients with advanced symptoms (Figure 6B).

**Figure 6.**
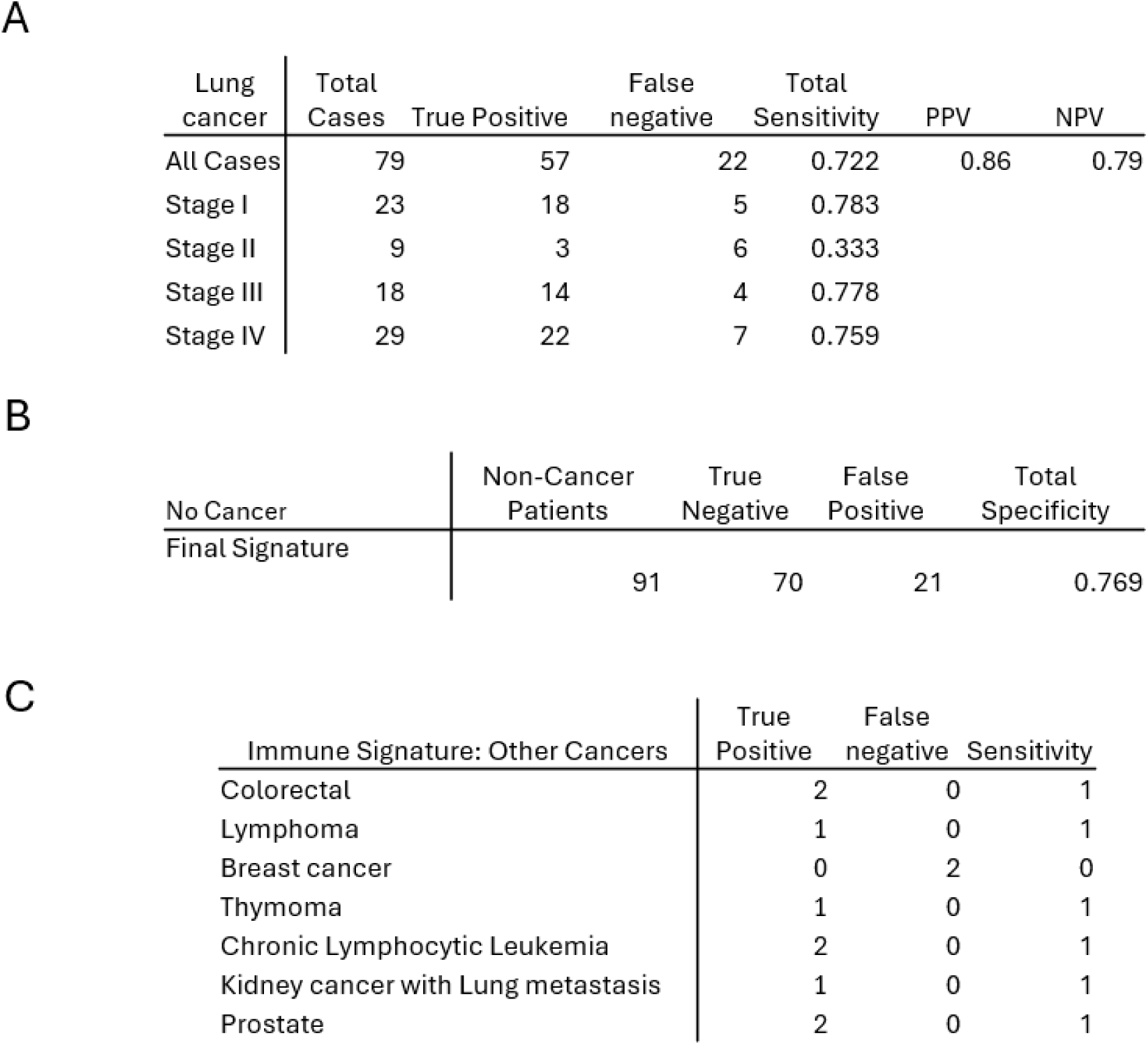
Tables of the performance of the final signature (the combined late-fusion model) when thresholding the risk score according to Youden’s criterion: sensitivity (A) and specificity (B). Table C shows the performance of the immune signature with those patients who were diagnosed with a non-lung cancer; the lung CT scan therefore not being relevant as a predictor of non-lung cancer.

Data were analysed for the 11 patients who received a non-lung cancer diagnosis. As the primary disease was not a lung cancer, the efficacy of the lung imaging data was unclear, and hence we used the immune signature alone. The sensitivity of this signature to detect cancer in these cohorts was 82%, with 9/11 cancers detected (Figure 6C). The two missed cancers were both breast cancer.

## Discussion

### Immune Subpopulations

We have previously shown that early on-treatment changes in peripheral levels of memory CD8 T-cells can predict response in patients with head and neck cancer ^30^, and that pre-treatment levels of a subpopulation antigen presenting capable myeloid cells can predict treatment outcome. We therefore postulated that that the systemic immune response may also be indicative of diagnosis of cancer in this study and highlight the fact that the signature obtained from a simple blood sample had a stronger predictive power than a chest CT scan. One of the strongest predictors in the signatures was the upregulation of KIR+ CD8 T lymphocytes. KIR3DL1 is part of the Killer Cell Inhibitory Receptor (KIR) family. Its expression on CD8 T cells has been linked to effector CD8 T cells with reduced proliferative capabilities and poor IFNg production following TCR engagement ^37, 38^. The inhibitory effect of KIR binding to HLA has been shown to not directly impact the T lymphocyte’s ability to degranulate but rather to compromise activation induced transcription which is required for clonal expansion and cytokine production ^39^. KIR expression has been linked to demethylation following chronic TCR stimulation which is consistent with cancer immunosurveillance ^40, 41^. In our data set, the KIR+ CD8 population exhibited a low level of expression of CD107a (Lamp1) indicating that those cells did not engage in degranulation which is even more indicative of their exhausted functional state (Supplementary Figure 5, Supplementary Figure 6).

Although KIR expression cannot not be, strictly, considered an exclusive feature of tumour specific T lymphocytes (e.g. there is a number of works describing its implications in viral immunology, especially HIV), there are works that demonstrated that KIR+ lymphocytes are restricted to specific tumour infiltrating lymphocytes subsets in renal cell carcinoma^42^ and melanoma^43^ and the phenotype of this cells closely resemble that of CD28 negative memory T cells. The latter work, actually, offers an intriguing hypothesis, claiming that the inhibitory nature of KIRs (they interfere with the interaction with HLA), serves the purpose of preventing the excessive cytotoxic activity of CTLs that bear a TCR specificity to self-antigens (peripheral tolerance), still maintaining reactivity against tumoral cells that might downregulate class I expression (a characteristic mechanism of tumour immune surveillance evasion).

Moreover, recent works suggests an active regulatory function of KIR+CD8+ lymphocytes that, for example, seems to suppress anti-tumour immunity in patients with melanoma^44^. This peculiar subset of CD8 T-lymphocytes resemble the phenotype of mouse Ly49+CD8+ Tregs^45, 46^ and appears to be particularly relevant in both auto-immune conditions, such as lupus and celiac disease in which they efficiently eliminate gliadin-specific CD4 T-lymphocytes, and SARS-CoV-2 ^47^, making them particularly relevant in the context of self-tolerance.

Of particular interest is that this signature could distinguish between a cancer and a non-cancer pathology. As most of the other pathologies were linked to an infection or an allergic reaction, the upregulation of cDC2 in those patients is only logical. cDC2 are a subtype of DCs that play a key role for antigen presentation for a type 2 helper response (TH2) ^48^ which is key in controlling humoral immunity. cDC2 have also been correlated with their ability to activate a TH17 response, which has been shown to be upregulated in chronic allergy ^49, 50^. The cDC2 cells expressed high levels of HLADR and CD38, indicating that they were indeed functional and pro-inflammatory. This subpopulation of myeloid cells did not express PDL2 indicating that it was not functionally suppressed (Supp. Fig. 6).

In recent years, the immune-surveillance theory and the efficacy of anticancer immunotherapies have been progressively more, and closely connected to the concept of immunometabolism and the microbiome profile of patients with cancer^51^, making clear that immune evasion (a contributing factor to tumorigenesis) may be affected directly by metabolic pathways e.g. lactate metabolism^52^, and these metabolic dysregulations can, in turn, affect immune cell function and tumour cell progression^53^. The KIR3DL1 CD8 T cells could represent a hypofunctional T cell subset^54^ that contributes to tumorigenesis.

It is now recognised that T and NK quiescence, activation, and exhaustion are regulated by cell metabolic states that work in conjunction with immunological stimuli^55^. In this perspective it would be of pivotal interest including in future models modalities that consider metabolomic parameters that could strengthen, because of causative correlations, our exquisitely immunological observations. The other consistent immunological parameter that links with the cancer pathology is reduced peripheral blood cDC2. Further work is required to investigate whether this is correlated with an increased degree of infiltration by cDC2 as previously reported in head and neck cancers^56^.

### Deep Learning Radiomics and Combined Model Generation

There has been significant recent activity in the development of techniques to extract novel information from medical images and other patient sample data and exploit it for clinical use^57^. Links between imaging and the immune system have also been made^58–62^. Grossman et al.^58^ evaluated radiomic texture and shape features in NSCLC and found correlations with immune response, inflammation and survival, and a combined signature that combined clinical, genetic and radiomics achieved a test set concordance index of 0.73 against OS.

Deep Learning networks have shown great promise in several fields, including cancer diagnosis, for extracting significant features from complex data, but their often black-box nature is one barrier to clinical use^63^. Autoencoders are able to learn semantically meaningful representations of images in an unsupervised manner and offer some advantages: training on an external data set and transparency through the exploration of the latent space parameters. Examples have been published in using epigenetic data to determine subtypes of lung cancer^64^, and image-based lesion detection and segmentation^65^.

The method for combining radiomics with blood data, needs consideration (i.e. data fusion^66^) as do the algorithms used to generate the predictive models. A model formed from many covariates can easily have exceptional performance on the training set, but this often does not translate to test and validation data sets (the problem of overfitting). Many studies use parameter regularisation such as LASSO or Elastic Net algorithms for signature generation and covariate selection. In particular, a study investigated the feasibility of preoperative 18F-fluorodeoxyglucose (FDG) PET/CT radiomics with machine learning (LASSO) to predict microsatellite instability (MSI) status in patients with colorectal cancer (CRC), with a test set performance (ROC AUC) of 0.867 from a published set of radiomics features^67^. Methods more advanced than regularisation offer a greater potential for generalisation to test and validation sets. For instance, neural networks and XGBoost models have achieved high ROC AUC of 0.75-0.8 in multimodal data sets^68–70^. Bayesian methods offer robust models that are generalisable and more interpretable and can have automatic feature selection that aid interpretability^71, 72^.

While a completely independent validation cohort with deep radiomic and immune profiles is currently outside the scope of this manuscript, we have repeated the randomisation of the patient cohort and machine learning 8 times, and the same two immune variables (reduced cDC2 levels and increased KIR3DL1+ CD8 T cells) have been retained in the model in the majority of these 8 independent model runs, lending credence to the selected predictive features.

A ROC AUC of 0.8 is within the realms of what may be clinically useful depending on the context. Choosing the optimal point on the ROC curve, to define operational sensitivity and specificity, may not be an obvious one for the clinical situation. Indeed, the multi-cancer Grail tests^21, 73^ have a similar AUC. The Galleri test has a specificity of around 99% but a sensitivity as low as 17% (depending on cancer and stage)^74^.

The 6mm clinical cutoff lesion size may limit the applicability of this model and we may obtain better results in early cancers is this is relaxed. This would however require a different benchmark to the optimal NOLCP pathway and longer follow-up to obtain definitive diagnosis which was not in the scope of this study. Further improvement may be obtained by retraining the deep learning autoencoder on a dataset with a greater match to the problem to avoid any domain mismatch between the LUNA16 dataset and the study acquired images. There may be other sources of bias in the training and test datasets that cannot be controlled for in this study, such as a lack of heterogeneity or cancer subtypes.

This present study has data collected in a prospective and standardised manner and used Bayesian machine learning techniques that emphasise reproducibility. Furthermore, ours is a “white box” method that generates an optimal set of parameters and weights that are available for interpretation. We explore a combination of deep learning radiomics and pre-determined standardized features in the form of the well-published filtration-histogram based texture analysis approach (using the commercially available TexRAD software) for feature selection from CT images.

## Data Availability

All data produced in the present study are available upon reasonable request to the authors from the DIO: 10.18742/28070390

## Contributors

Contributions to this manuscript are as follows: Data curation: RM, BG, SE, LD, MT, XJ, CS, SL, EC, AT, LH, RE, RS, DW, SC, PB; Formal analysis: RM, BG, SE, EC, AC, PB; Investigation: RM, SE, LD, XJ, CS, SC, PB; Methodology: RM, BG, SE, LD, XJ, CS, AC, JS, TN, PB; Resources: RM, LD, KD, SL; Validation: RM, LD, MR, AC, TN, PRB; Visualisation: RM, PB; Software: SE, MT, MR, AC, PB; Supervision: BG, AC, AG, JS, TW, PB, TN; Writing – original draft: RM, BG, SE, KD, JAS, TW, PB, TN; Writing – review & editing: LD, PB, TN; Conceptualisation: AG, JS, TW, TN; Funding acquisition: AG, JS, TW, TN; Project administration: TN. All authors read and approve the final version of the manuscript. RM and PB had access to the underlying data for the multivariate analysis.

## Data Sharing Statement

Model generation was performed using the MsaWrapper R package (https://github.com/paulbarber/Lung_EDx_Manuscript_Processing and https://github.com/paulbarber/msaWrapper) and Saddle Point Signature (commercial software distributed by Saddle Point Science Ltd).

Anonymised data used for this analysis is available at reasonable request (to comply with patient consent) from: kcl.figshare.com/account/articles/28070390

## Acknowledgements

We are grateful to all participating staff at The Lister, Hertford County, UK and New QEII Hospitals, Welwyn Garden City, UK who were responsible for patient recruitment and sample collection.

This work was funded by the CRUK grant [C1519/A27375]. SE, EC and JAS were further supported by core funding from the Wellcome Trust/EPSRC Centre for Medical Engineering [WT203148/Z/16/Z] and by the National Institute for Health and Care Research (NIHR) Clinical Research Facility at Guy’s and St Thomas’ NHS Foundation Trust. The views expressed are those of the author(s) and not necessarily those of the NHS, the NIHR or the Department of Health and Social Care. Work was undertaken at UCLH/UCL, which received a proportion of the funding from the UK’s Department of Health’s NIHR Biomedical Research Centre’s funding scheme.

