## Supplementary Information for "Fusing Data from CT Deep Learning, CT Radiomics and Peripheral Blood Immune profiles to Diagnose Lung Cancer in a Cohort of Patients Experiencing Symptoms"

**Mustapha et al.**

**Supplementary Information**

### Methods

#### CT scans

Patients with a clinical or radiological suspicion of lung malignancy underwent thoracic CT scans at a single hospital site. Different CT scanners were used to acquire the images and scan parameters varied according to patient habitus, but the scans were all performed to a diagnostic standard which included 1mm slice thickness and the administration of intravenous contrast unless contraindicated.

The scans were pseudo-anonymized and reviewed on a proprietary workstation by a research radiographer with 26 years of cross-sectional imaging experience in oncology. The radiographer was blinded to each patient's medical record and any related clinical opinion. They identified post-contrast scans that contained a suspicious lung lesion greater than 6mm in the axial dimension and excluded the remaining scans from further analysis. For each scan, a single axial image demonstrating the lesion at its largest was selected and uploaded to a proprietary software for textural analysis.

A second research radiographer with 9 years' experience in image segmentation manually drew a region of interest around the lesion on each pre-selected image, carefully excluding adjacent soft tissue structures considered normal anatomy. The software applied an automatic thresholding algorithm to exclude air (<-50 HU) and calcification (>200 HU) and used a filtration-histogram technique to enhance and extract imaging features of varying sizes and intensity changes. See Supplementary Figure 1.

#### CT Texture Analysis (CTTA)

CT Texture Analysis (CTTA) was performed using a commercially available research software called TexRAD Research (Feedback Medical Ltd., London/Cambridge, UK). Furthermore, a version of the software called TexRAD Lung had CE mark for potential use in lung cancer on PET/CT. The commercially available research software has an embedded DICOM viewer and is installed as a server-web-client solution within the hospital network, communicating via DICOM protocol with the PACS or data-archive (mini-PACS) to receive DICOM images for analysis. The following process was undertaken exclusively within the TexRAD Research software.

CT Texture Analysis (CTTA) comprised a filtration-histogram based technique, to extract and enhance features of different sizes and intensity-variation corresponding to the spatial scale filter (SSF), which varied from SSF=0,2,3,4,5,6 mm (object-size in radius). Here, SSF=2 corresponded to fine texture, SSF=3-5mm corresponded to medium texture and SSF=6mm corresponded to coarse texture, where SSF=0 corresponded to no filtration. Quantification of texture using statistical and histogram-based techniques included the following metrics: mean intensity (mean), standard deviation (sd), entropy, mean of positive pixels (mpp), skewness and kurtosis. Texture was quantified within each filtered (SSF=2-6) map and on the conventional CT image without filtration (SSF=0). A CT phantom simulation study described how the filtration-histogram based texture features reflected different components of tissue/tumour heterogeneity in terms of object size, number (concentration), density/intensity in relation to the background tumour/tissue parenchyma [1-3]

The filtration-histogram CTTA technique visually and quantitatively reflects tumour heterogeneity, which is essential for translating such techniques from research to clinic. Other radiomic techniques/pipelines (e.g. PyRadiomics) looks at numerous metrics using higher order statistics which make the approach abstract and less biologically intuitive and increases false discovery rate. Furthermore, the filtration-histogram based CTTA technique has undergone extensive robust qualification process in terms of technical validation (test-retest, reliability/reproducibility[4], phantom studies[1], different scanning acquisition protocols and multi-centre evaluation[5, 6]), biological rationale (correlation with stage, severity[7], histology[8], immunohistochemistry – hypoxia, angiogenesis markers[9] etc., molecular-signatures/genetic-mutations – KRAS[10], EGFR[11] etc.), key clinical utility and validation (prognostication[5, 12], risk for nodal-metastasis[13], response to conventional[14], novel targeted[15], immunotherapies[16-18] from single and multi-centre studies[5]), clinical-adoption[19], barriers to clinical translation[20], and cost-effectiveness[21]. An example can be seen in Supplementary Figure 2.

#### Deep Learning Autoencoder (DLA)

Deep learning (DL) unsupervised autoencoder (AE) models were trained on a publicly available lung CT dataset, LUNA 16 [22]. No AE training was performed with the data set of this present study. Three network architectures were tried:

**GAN-AE:** A generator network was first trained to produce lung lesion CT images from normally distributed random latent vectors using the generative adversarial network (GAN) framework[23]. Subsequently, the generator was used as the decoder of an autoencoder network, with the decoder frozen and the encoder trained with an L1 pixel-space loss and a weighted perceptual loss using features computed with the final GAN discriminator. This approach is conceptually similar to, but a simpler version of, other methods in the literature such adversarial autoencoders [https://arxiv.org/pdf/1511.05644]. GAN-AE models were parameterised by the latent space size, $n \in \{32,64,128,256,512\}$, and the weighting of the perceptual loss, $\text{lambda} \in \{0,0.1,0.5,1,2,5\}$.

**Beta-VAE:** A Variational AE [24] trained with a combination of L1 pixel-space reconstruction loss and Kullback-Leibler (KL) divergence, with a variable weighting on the KL-divergence loss. beta-VAE models were parameterised by the latent space size, $n \in \{32,64,128,256,512\}$, and KL-divergence weighting, $\text{beta} \in \{1\times10^{-5}, n\times10^{-6} | n \in \{1,2,3,\ldots,9\}\}$.)

**Beta-VSSAE:** A variational scale-space AE [25] with a variable weighting on the KL-divergence loss, $\text{beta} \in \{1\times10^{-5}, n\times10^{-6} | n \in \{1,2,3,\ldots,9\}\}$. The beta-VSSAE encodes the image at multiple scales, with the latent space sizes determined by a scaling factor $n\_mult \in \{1,2,4,8\}$. This factor is multiplied by the image size at each scale level to compute the corresponding latent space size.

All images used with the AE models were resampled to 0.625mm isotropic and normalised between -1 and 1, then cropped to a central patch of 64x64. The models were trained with Adam optimiser, with a learning rate of 0.0001 and batch size 64. During training, data augmentation was applied in the form of horizontal and vertical flips, Gaussian blurring, rotation, scaling and translation.

Following training of the AE models, latent parameters, after the encoder stage, were taken forward to multi-modal data fusion since they should contain sufficient information to recreate all the major features of the images. Latent feature interpretation could be performed using the decoder stage. By feeding latent variables into the decoder and changing latent values, one-by-one, the effect on the image can be observed.

After the initial training and preliminary evaluation of the networks, a GAN model with 32 latent parameters, lambda = 0.5 (GAN32_0.5), was taken forward to data fusion because of its good performance and good interpretability. Models with higher numbers of latent parameters often produced parameters that were not visibly interpretable.

#### Peripheral Blood Sampling and Processing.

Blood was collected from patients at the time of their CT scan. 10ml of venous blood were collected into SST vacutainers for downstream exosome isolation (Becton Dickinson, UK 367954). 12 ml venous blood were collected into K2-EDTA tubes for downstream PBMC isolation (Becton Dickinson, UK 367873). Samples were stored at 4 degrees for no more than 24 hours before being processed. Plasma was isolated from SST tubes following centrifugation at 1000g for 10 minutes at 4°C then frozen at -80°C. PBMCs were isolated from whole blood using differential centrifugation. Briefly the whole blood was diluted 1:1 in PBS. The blood was then layered over ficol and spun without brakes at 400g for 40 minutes. The buffy coat containing the PBMCs was then collected and washed twice with PBS before being frozen down in FBS 10% DMSO by controlled freezing.

#### Flow Cytometry

Frozen PBMC samples were thawed and stained with Fixable viability dye (Zombie UV, Biolegend) followed by two different panels of membrane markers: a lymphoid staining panel (panel L) and a myeloid staining panel (panel M). Two different panels full list of both antibody panels in Supplementary Table S1, each antibody was validated and titrated (column "µl/$10^6$ cells") in preliminary experiments involving PBMC samples obtained from healthy donors, eventually activated with different means to simulate specific immunological scenarios such as lymphocytes activation or myeloid expansion (e.g. phytohemagglutinin or cytokines such as GM-CSF and IL-4). Patients' samples were acquired in a Fortessa II flow cytometer (BD, Berkshire, UK) via the use of baseline settings and calibration beads (BD FACSDiva™ CS&T Research Beads, BD) and analysed with a custom R script. The script uses a combination of supervised and unsupervised methodology and can be divided into 3 stages. Real viable events are defined by performing clustering based on the forward scatter (area and width) side scatter (area and width) and viability staining. Those clusters that satisfy set thresholds for live viable cells are now clustered in an unsupervised approach into 364 populations per panel using the flowSOM package (Bioconducter). Those clusters are then aggregated into predefined super populations as per the Supplementary Table S2. Using these definitions, the distribution of the predefined super populations per patient were determined. Populations were excluded if the population was not present in the majority of patients. The percentages of the remaining super-populations were included in the risk signature generation.

#### Exosome Analysis by Dotblot

##### Isolation and characterisation of exosomes

Exosomes were isolated from frozen plasma samples using ultracentrifugation. After thawing, plasma was centrifuged at 10,000 x g to remove the platelets. The supernatant was collected and diluted 1:1 with 0.1µm filtered PBS (fPBS) and centrifuged twice at 12,200x g at 4°C for 25 minutes to remove larger vesicles from the plasma. The supernatant was centrifuged at 100,000 x g for 2 hours, 4°C in a OptimaTM MAX-XP Ultrancentrifuge equipped with a TLA-55 rotor (Beckman Coulter, Brea, California, USA). The exosome pellet was washed in fPBS and ultracentrifuged at the same speed for 1 hour, 4°C. The final exosome pellet was resuspended in fPBS and stored at -80°C. Nanoparticle tracking analysis (NTA) was performed using the Nanosight LM10 system (Malvern Panalytical) equipped with a LM15 laser unit to measure the size and concentration of the exosomes. All samples were diluted appropriately in 1ml fPBS and injected into the instrument using a syringe. Each sample was counted in replicates of 5, collecting 30 second videos per sample. The camera level used remained the same for each sample to allow for comparison of particle counts and fPBS was also run as a control. For all experiments the camera level used was 12 and the detection threshold used for analysis was set to 2.

#### Protein Detection using Immuno-blot

Exosome suspensions were blotted onto a 0.45µm nitrocellulose membrane (AmershamTM, Germany) by suction through 96-well Bio-Dot® microfiltration apparatus (Bio-Rad, Hercules, CA, USA). The membrane was blocked with 5% non-fat milk in tris-buffered saline with 0.05% Tween-20 (TBST) for 1 hour at room temperature. After Three washes with TBST (TBS with 0.05% Tween-20), membranes were incubated overnight at 4 °C with an appropriate concentration of primary antibody (supplementary table 3). After washing three times with TBST, the membranes were incubated with secondary antibodies, IRDye ® 680RD goat anti-rabbit IgG and 800CW goat anti-mouse IgG (LiCOR Biosciences, Lincoln, NE, USA) for 1 hour at room temperature and visualised using Odyssey CLx Imager (LiCOR Biosciences, Lincoln, NE, USA) See Table S3 for a list of the antibodies used. Images were quantified using Image StudioTM (version 3.1). Protein expression was normalised to ALIX protein expression.

#### Training and Test Set Formation

The whole data set was split into training and test sets in the proportion 3/4 to 1/4 respectively, using the R MatchIt package to ensure balanced cohorts. This splitting was repeated to gather statistics on the training and test set prediction performance.

#### Multivariate Signature Generation

Covariate reduction prior to model training was performed to exclude highly mutually correlative variables and to exclude those that had a very low correlation with outcome class. Correlative covariates from each modality were removed by detecting pairs with a Kendall rank correlation >0.5 and removing the covariate of the pair with the lower Kendall correlation with outcome. The autoencoder latent covariates were also excluded if the Kendall correlation with outcome was <0.15.

Data fusion was performed using three methods (Figure 1). Early fusion simply concatenated the tables of reduced covariates to form a new set encompassing all modalities. Intermediate fusion used the covariates selected for the models of each modality to form a new data set. Late fusion combined the risk scores generated from each unimodal model in a new model.

The generation of predictive signatures was performed by Bayesian multivariate regression (BMR) with repeated cross-validation and backwards elimination with the aim of reducing overfitting using the "batch regression" option of the Saddle Point Signature software (Saddle Point Science Ltd., London, UK) according to methods that were previously published. [26-28]

Covariates were normalized to zero mean and unit standard deviation such that the importance and significance of covariates can be judged by their assigned beta value (β) in the proportional hazards model, and corresponding hazard ratio (HR) equal to $e^{2\beta}$. A negative β value reflects a lower risk is associated with that covariate.

The Saddle Point Signature software additionally judges the performance of similar randomized data, which most often has β values around zero and within a critical range, such that any real covariate that has a β value outside this critical range can be judged to be performing significantly better than randomized data.

A Risk Score should be generated by combining covariate raw values in a linear model according to their given weights and intercept. The calculation could thus be performed in a suitable spreadsheet program by someone with no programming experience. Future application of the risk signature should replace any missing or poor quality values with a representative average value (that should be determined in future studies of larger patient sets).

Final models were trained on all data using the covariates which appeared in 50% of train/test split runs. With these reduced sets of covariates, overfitting on new data should be minimised, and the repeated test set performances give an indicator of future performance.

#### Supplementary Figures and Tables

| Target | Conjugation | Clone | Ref | ul/1x10$^6$ cells |
|---|---|---|---|---|
| CD19 | BUV395 | HIB19 | 740287 | 2 |
| CD15 | BUV395 | W6D3 | 740318 | 2 |
| CD33 | BUV395 | WM53 | 740293 | 2 |
| CD20 | BUV395 | 2H7 | 563782 | 2 |
| CD14 | BUV395 | MφP9 | 563561 | 2 |
| CD34 | BUV395 | 581 | 563778 | 2 |
| CD56 | BUV737 | NCAM16.2 | 612766 | 3 |
| CCR7 | BV421 | G043H7 | 353208 | 3 |
| CD4 | BV510 | SK3 | 344634 | 2 |
| CD11c | BV605 | 3.9 | 301636 | 2 |
| CD25 | BV650 | M-A251 | 563719 | 3 |
| CD25 | BV650 | 2A3 | 740634 | 3 |
| CD16 | BV711 | 3G8 | 563127 | 2 |
| CD107a | BV785 | H4A3 | 328644 | 3 |

| | | | | |
|---|---|---|---|---|
| CD8 | FITC | RPA-T8 | 301050 | 4 |
| CD3 | PerCP/Cy5.5 | UCHT1 | 300430 | 3 |
| KIR3DL1 | PE | DX9 | 312708 | 3 |
| CD45Ro | PE/Cy7 | UCHL1 | 304230 | 3 |
| CD127 | AF647 | HIL-7R-M21 | 558598 | 10 |
| CD127 | APC | eBioRDR5 | 17-1278-42 | 4 |
| TCRgd | APC-R700 | 11F2 | 657706 | 4 |
| HLADR | APC/Fire750 | L243 | 980412 | 4 |
| CD3 | BUV395 | UCHT1 | 563546 | 3 |
| CD56 | BUV395 | NCAM16.2 | 563554 | 4 |
| CD15 | BUV737 | W6D3 | 741876 | 2 |
| CD24 | BV421 | ML5 | 311122 | 2 |
| HLADr | BV510 | L243 | 307646 | 5 |
| PDL2 | BV605 | MIH18 | 742718 | 3 |
| CD38 | BV650 | HB-7 | 356620 | 2 |
| CD11c | BV711 | S-HCL-3 | 744438 | 3 |
| CD27 | BV786 | M-T271 | 740972 | 4 |
| CD16 | FITC | CB16 | 11-0168-42 | 2 |
| CD19 | PerCP/Cy5.5 | HIB19 | 302230 | 3 |
| CD66b | PE | G10F5 | 305106 | 2 |
| IgD | PE/Dazzle594 | IA6-2 | 348240 | 4 |
| CD33 | PE/Cy7 | P67.6 | 366618 | 2 |
| CD14 | APC | HCD14 | 325608 | 6 |
| CD11b | APC/Cy7 | ICRF44 | 301342 | 2 |

Supplementary Table S1. List of antibodies, used in flow cytometry.

| | Dump | CD19 | IgD | CD27 | CD38 | CD24 | CD33 | CD14 | CD11b | CD11c | CD15 | CD66b | CD16 | HLADr | PDL2 |
|---|---|---|---|---|---|---|---|---|---|---|---|---|---|---|---|
| Total B-lymphocytes | - | + | | | | | - | | | | - | | | | |
| Memory pre-switch B-lymphocytes | - | + | + | + | | | - | | | | - | | | | |
| Memory post-switch B-lymphocytes | - | + | - | + | | | - | | | | - | | | | |
| DN B-lymphocytes | - | + | - | - | | | - | | | | - | | | | |
| Naive B-lymphocytes | - | + | + | - | | | - | | | | - | | | | |
| Transitional B-lymphocytes | - | + | - | - | + | + | - | | | | - | | | | |
| Mature naive B-lymphocytes | - | + | - | + | - | - | - | | | | - | | | | |
| Plasma B-lymphocytes | - | + | - | + | + | - | - | | | | - | | | | |
| Myeloid cells total | - | - | | | | | + | | | | | | | | |
| PMN total | - | - | | | | | | | | | + | | | | |
| Classical monocytes | - | - | | | | | + | + | | | - | | - | + | |
| Intermediate monocytes | - | - | | | | | + | + | | | - | | + | + | |
| Non-classical monocytes | - | - | | | | | + | - | | | - | | + | + | |
| MO-MDSC | - | - | | | | | + | + | + | | - | | | - | |
| Im-myeloid | - | - | | | | | + | - | + | | - | | | - | |
| PMN-MDSC | - | - | | | | | - | - | + | | + | | | | |
| Total DC | - | - | | | | | + | | | | - | | | + | |
| cDC2 | - | - | | | | | + | - | - | + | - | | - | + | |
| CD16-DC | - | - | | | | | + | - | - | + | - | | + | + | |
| MO-DC | - | - | | | | | + | + | | + | - | | | + | |
| CD38+ Classical Monocytes | - | - | | | + | | + | + | | | - | | - | + | |
| CD38- Classical Monocytes | - | - | | | - | | + | + | | | - | | - | + | |
| PDL2+ myeloid cells | - | - | | | | | + | | | | | | | | + |
| PDL2+ B-lymphocytes | - | + | | | | | - | | | | - | | | | + |
| DUMP- unidentified | - | - | | | | | - | | | | - | | | | |
| Lin- HLADR+ | - | - | | | | | - | | | | - | | | + | |
| CD3+CD33+ | + | | | | | | + | | | | | | | | |

| | Dump | CD3 | CD4 | CD8 | TCRgd | CD25 | CD127 | CCR7 | CD45Ro | CD56 | CD16 | KIR3DL1 | CD11c | CD107a | HLADR |
|---|---|---|---|---|---|---|---|---|---|---|---|---|---|---|---|
| Total T-lymphocytes | - | + | | | | | | | | | | | | | |
| Total CD4 T-lymphocytes | - | + | + | | - | | | | | | | | | | |
| CD4+ T-regs | - | + | + | | | + | - | | | | | | | | |
| CD4+ Effector | - | + | + | | | | | - | - | | | | | | |
| CD4+ Naive | - | + | + | | | | | + | - | | | | | | |
| CD4+ Central memory | - | + | + | | | | | - | + | | | | | | |
| CD4+ Effector memory | - | + | + | | | | | + | + | | | | | | |
| Total CD8 T-lymphocytes | - | + | | + | - | | | | | | | | | | |
| KIR3DL1+ CD8 T-lymphocytes | - | + | | + | | | | | | | | + | | | |
| CD8+ Effector | - | + | | + | | | | - | - | | | | | | |
| CD8+ Naive | - | + | | + | | | | + | - | | | | | | |
| CD8+ Central memory | - | + | | + | | | | - | + | | | | | | |
| CD8+ Effector memory | - | + | | + | | | | + | + | | | | | | |
| Total TCRgd T-lymphocytes | - | + | | | + | | | | | | | | | | |
| KIR3DL1+ TCRgd T-lymphocytes | - | + | | | + | | | | | | | + | | | |
| TCRgd T-lymphocytes CD45RO+ | - | + | | | + | | | - | + | | | | | | |
| TCRgd T-lymphocytes CD45RO- | - | + | | | + | | | + | - | | | | | | |
| NK cells | - | - | | | | | | | | + | | | | | |
| NK CD16+ | - | - | | | | | | | | + | + | | | | |
| NK CD16- | - | - | | | | | | | | + | - | | | | |
| KIR3DL1+ NK cells | - | - | | | | | | | | + | | + | | | |
| NKT cells | - | + | - | - | - | | | | | + | | | | | |
| ILC cells | - | - | - | - | | | + | | | - | | | - | | |
| DUMP- undefined | - | - | - | - | - | | | | | - | | | | | |

Supplementary Table S2. Table showing the definitions for the super populations that were included in the risk signature generation.

| Antibody | Species | Clone | Isotype | Concentration | Company | Cat Number |
|---|---|---|---|---|---|---|
| Alix | Ms | 3A9 | Mouse IgG1 | 0.5mg/ml | Biolegend | 634502 |
| S100A9 | Rb | D5O6O | Rabbit IgG | 4.3mg/ml | Cell Signalling Technology | 34425 |
| P4AH1 | Rb | | Rabbit IgG | 0.3mg/ml | Proteintech | 12658-1-AP |
| TIGIT | Ms | A15153G | Mouse IgG2a | 0.5mg/ml | Biolegend | 372702 |
| PDL1 | Ms | MIH2 | Mouse IgG1 | 0.5mg/ml | Biolegend | 393602 |
| CD63 | Rb | EPR5702 | Rabbit IgG | 0.384mg/ml | Abcam | AB134045 |
| CTLA4 | Ms | 14D3 | Mouse IgG2a | 0.5mg/ml | eBioscience | 14-1529-82 |
| BAG6 | Rb | EPR9223 | Rabbit IgG | 0.347mg/ml | Abcam | AB137076 |
| LAG3 | Ms | 11C3C65 | Mouse IgG1 | 0.5mg/ml | Biolegend | 369302 |
| HER2 | Ms | 24D2 | Mouse IgG1 | 0.5mg/ml | Biolegend | 324402 |
| CD73 | Ms | AD2 | Mouse IgG1 | 0.5mg/ml | Biolegend | 344002 |
| EGFR | Ms | AY13 | Mouse IgG1 | 0.5mg/ml | Biolegend | 352902 |
| PD1 | Ms | EH12.2H7 | Mouse IgG1 | 0.5mg/ml | Biolegend | 329902 |
| CD226 | Ms | 11A8 | Mouse IgG1 | 0.5mg/ml | Biolegend | 338302 |
| ICOSL | Ms | 2D3 | Mouse IgG2b | 0.5mg/ml | Biolegend | 309402 |
| HER3 | Rb | D22C5 | Rabbit IgG | | Cell Singalling Technology | 12708 |
| B2M | Ms | 2M2 | Mouse IgG1 | 0.5mg/ml | Biolegend | 316302 |
| MET | Rb | | Rabbit IgG | | Cell Singalling Technology | 4560S |
| HLADR | Ms | L243 | Mouse IgG2a | 0.5mg/ml | Biolegend | 307651 |
| Alix | Rb | | Rabbit IgG | 0.5mg/ml | Abcam | AB225555 |
| CD63 | Ms | MEM-259 | Mouse IgG1 | 0.5mg/ml | GeneTex | GTX28219 |
| Mouse IgG1 | | MG-1-45 | | 0.5mg/ml | Biolegend | 401402 |
| Mouse IgG2a | | MG2a-53 | | 0.5mg/ml | Biolegend | 401502 |
| Mouse IgG2b | | MG2b-57 | | 0.5mg/ml | Biolegend | 401202 |
| Rabbit IgG | | EPR25A | | 1.675mg/ml | Abcam | ab172730 |

Supplementary Table S3 showing the antibodies using for exosome analysis by dotblots

| Cancer type | Number of patients | Average Age Bracket | Sex | | Smoking status | | | |
|---|---|---|---|---|---|---|---|---|
| | | | Male | Female | Ex-Smoker | Non-Smoker | Smoker | Unknown |
| Colorectal | 2 | 80-89 | 1 | 1 | | | | |
| Lymphoma | 2 | 60-69 | | | 2 | | | |
| Breast cancer | 2 | 70-79 | 1 | 1 | | | | 2 |
| Thymoma | 1 | 60-69 | | 1 | 1 | | | |
| Chronic Lymphocytic Leukemia | 2 | 80-89 | | 2 | | 1 | | 1 |
| Kidney cancer with Lung metastasis | 1 | 70-79 | 1 | | 1 | | | |
| Prostate | 1 | 60-69 | 1 | | | 1 | | |
| Melanoma | 1 | 60-69 | | 1 | 1 | | | |

Supplementary Table S4 showing characteristics of non-lung cancer patients. The 10-year age bracket of the average age is shown.

1

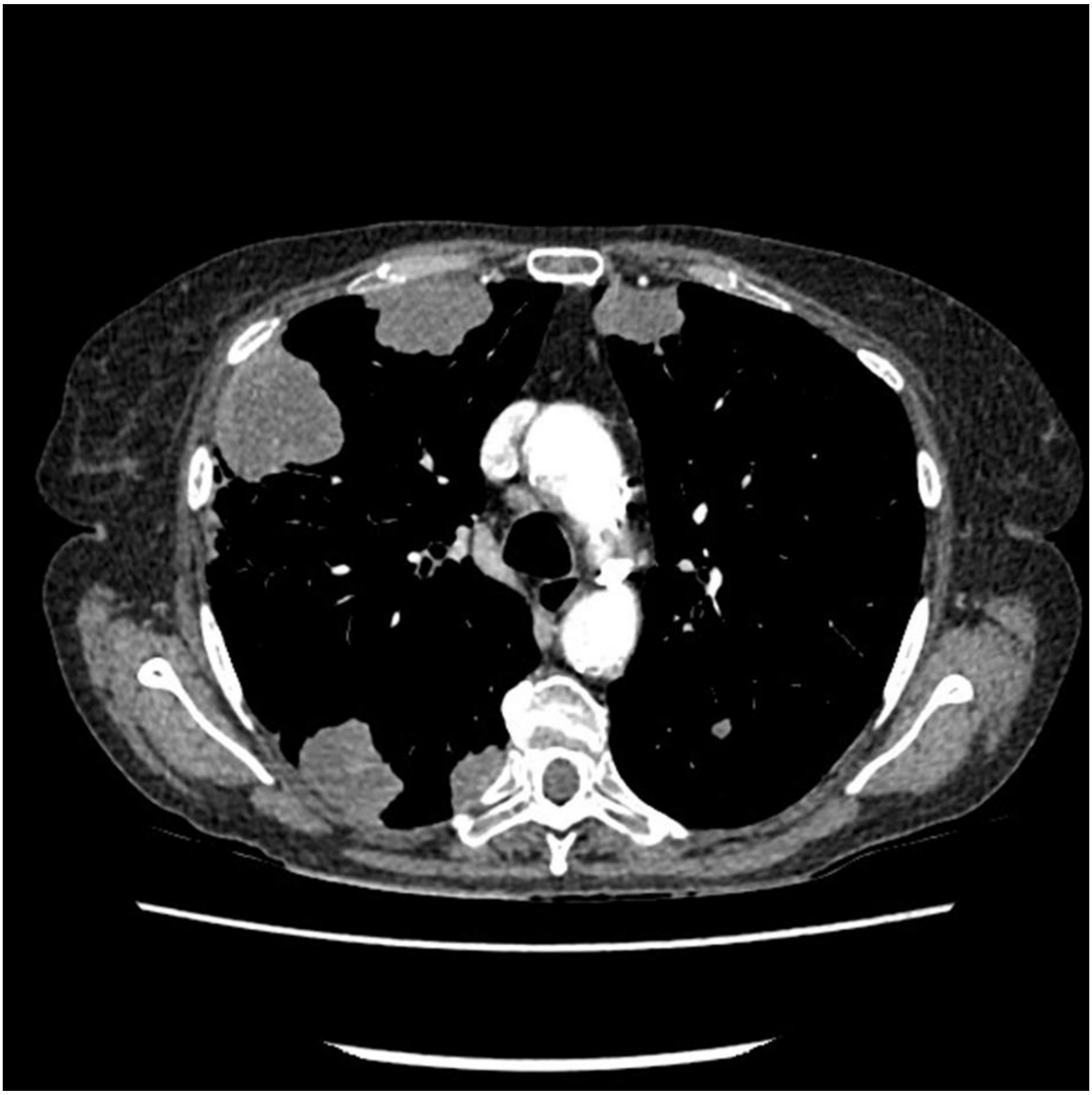

2

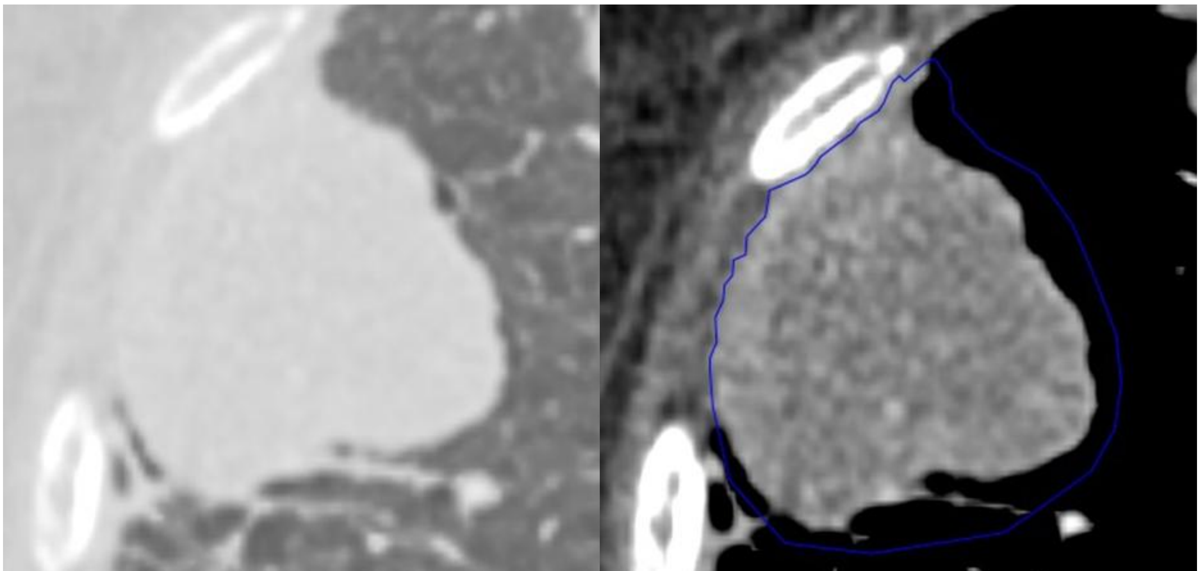

Supplementary Figure 1: Example of lesion identification from a CT scan and manually drawn region of interest. Top: Full lung cross-section. Bottom-left: Extracted lesion image. Bottom-right: Lesion image with initial manually drawn region of interest and automatic thresholding applied to the ROI.

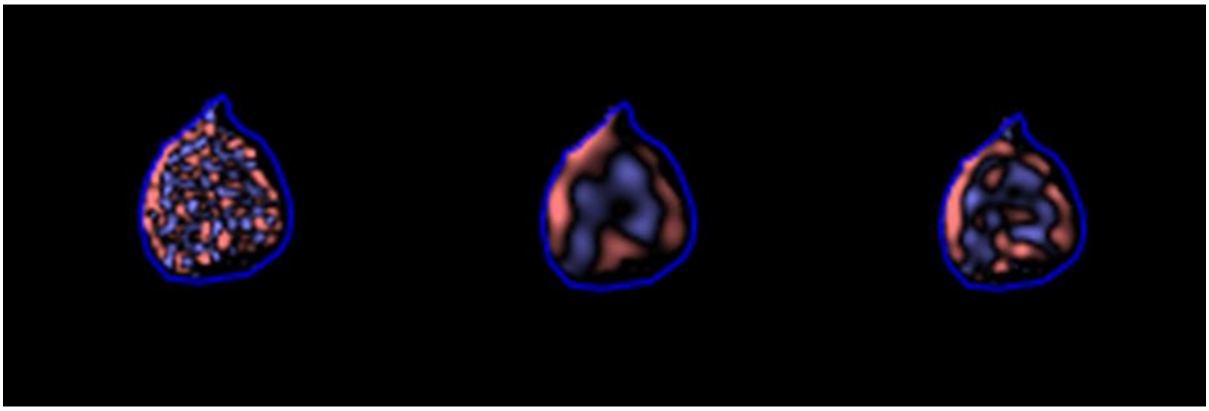

Supplementary Figure 2: Examples of the filtered texture maps as applied by the CTTA algorithm on the region of interest at different texture scales (Spatial scale filter - SSF & pink features reflects positive filtered intensity pixel values and blue features reflects negative filtered intensity pixel values). Left: fine (SSF=2mm), Middle: medium (SSF=4mm) and Right: coarse (SSF=6mm).

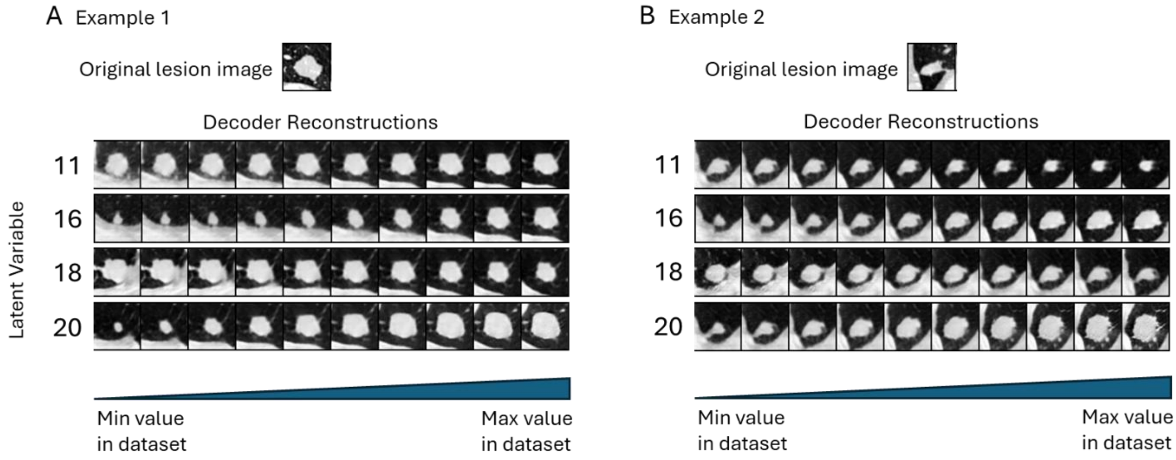

Supplementary Figure 3: Varying the value of significant DL latent variables fed into the decoder gives a visual interpretation of their effect. A and B show example lesion images and the effect of varying the latent variables from the minimum value found across all images in the dataset to the maximum value. It can be seen that variables numbered 16 and 20 seems to encode lesion size, and other variables encode combinations of pleural attachment and the morphology of the lesion and surrounding tissue.

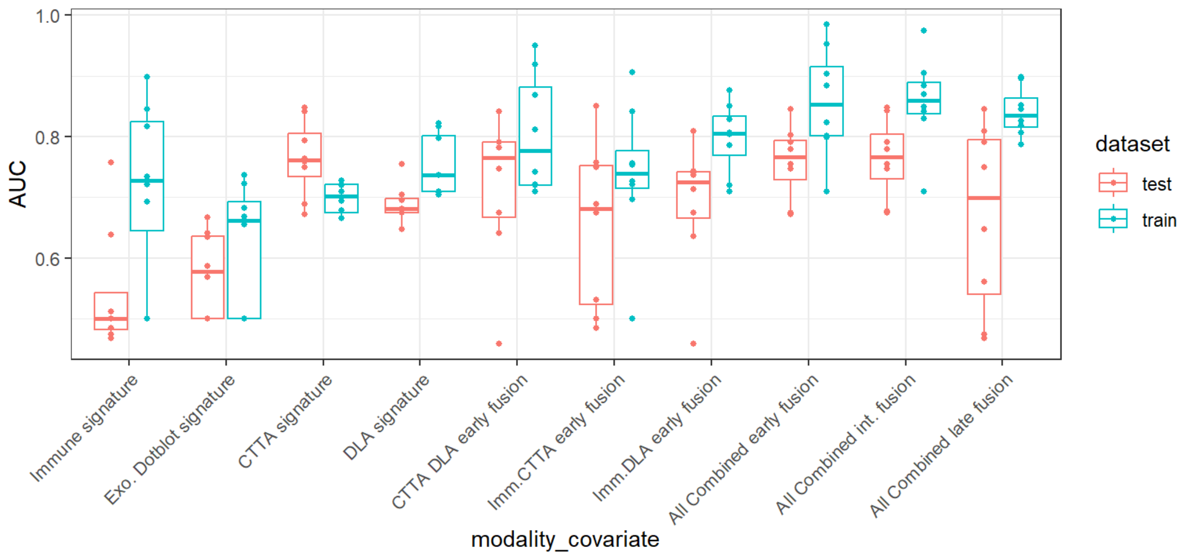

Supplementary Figure 4: Risk signature generation from repeated train/test splits. Summary of ROC AUC values on the training set (blue) and test set (red) for all the generated signatures using Elastic Net.

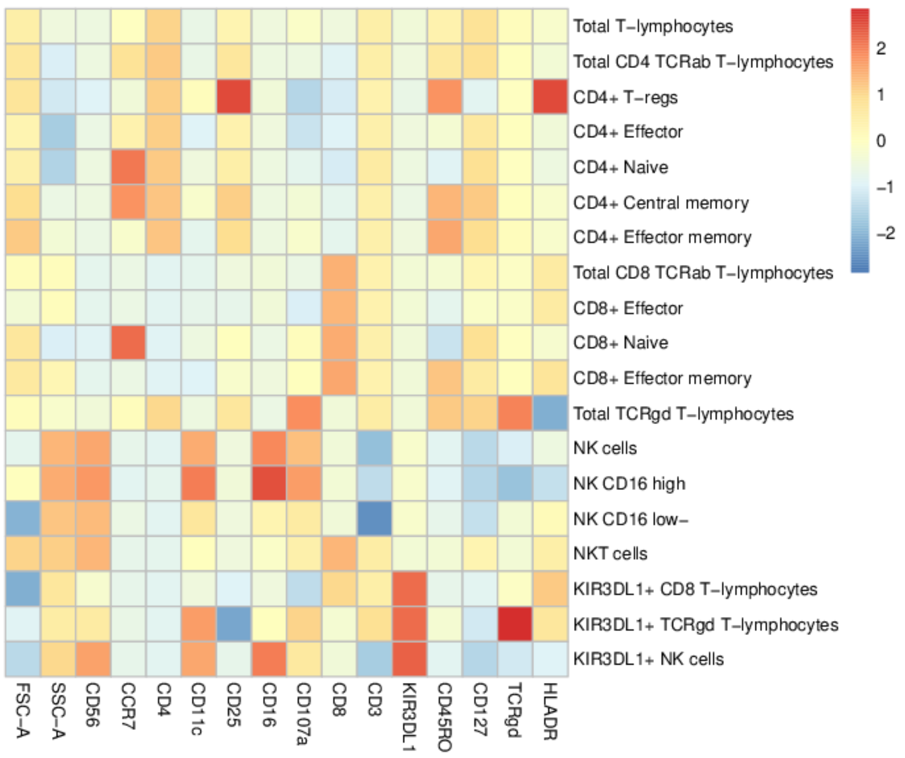

Supplementary Figure 5: Z-score MFIs of both morphological (FSC and SSC) and immune markers included in the lymphoid staining panel (panel L) for each super-cluster. The determination was conducted on an artificial sample created by sampling the same number of events from each patient.

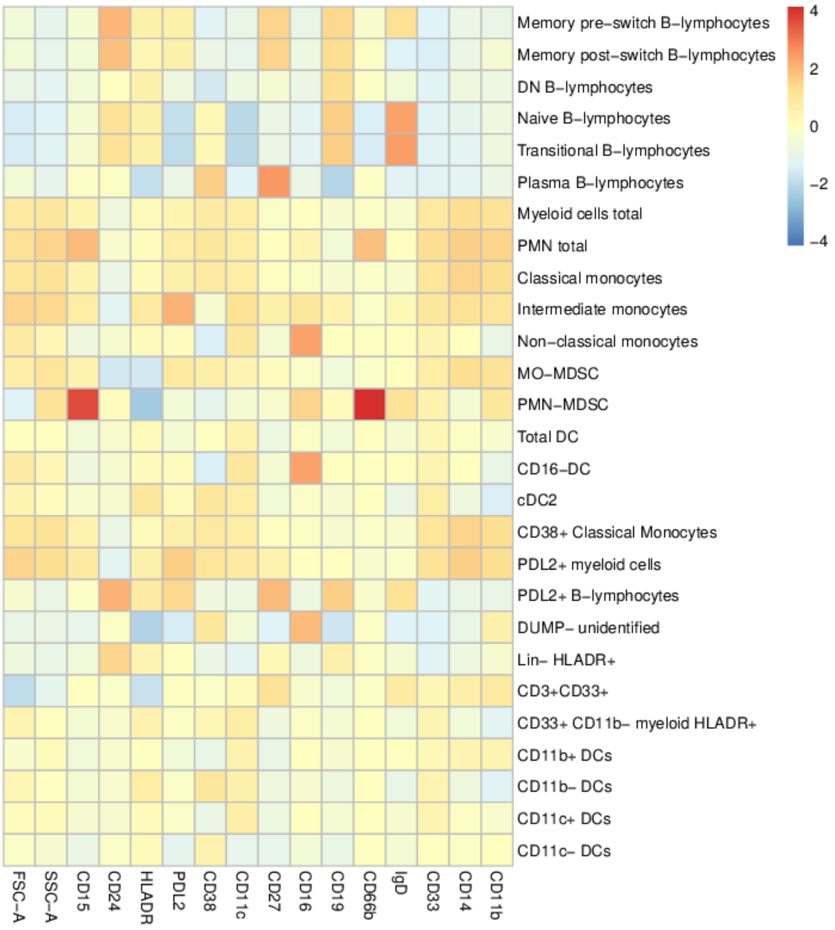

Supplementary Figure 6: same analysis as presented in supplementary figure 5, but pertinent to the myeloid staining panel (M).

16 Ravanelli M, Agazzi GM, Milanese G et al. Prognostic and predictive value of histogram analysis in patients with non-small cell lung cancer refractory to

platinum treated by nivolumab: A multicentre retrospective study. Eur J Radiol 2019; 118.
17 Ladwa R, Roberts KE, O'Leary C et al. Computed tomography texture analysis of response to second-line nivolumab in metastatic non-small cell lung cancer. Lung Cancer Manag 2020; 9 (3).
18 Zerunian M, Caruso D, Zucchelli A et al. CT based radiomic approach on first line pembrolizumab in lung cancer. Sci Rep 2021; 11 (1).
19 Miles KA. How to use CT texture analysis for prognostication of non-small cell lung cancer. . Cancer Imaging 2016.
20 Miles KA, Squires J, Murphy M. Radiologist Engagement as a Potential Barrier to the Clinical Translation of Quantitative Imaging for the Assessment of Tumor Heterogeneity. . Acad Radiol 2018; 25 (7).
21 Miles KA, Ganeshan B. Selection of patients with advanced non-small cell lung cancer for chemotherapy: Potential cost-effectiveness of CT texture analysis. *ECR*. 2012.
22 Setio AAA, Traverso A, de Bel T et al. Validation, comparison, and combination of algorithms for automatic detection of pulmonary nodules in computed tomography images: The LUNA16 challenge. Med Image Anal 2017; 42: 1-13.
23 Ian Goodfellow JP-A, Mehdi Mirza, Bing Xu, David Warde-Farley, Sherjil Ozair, Aaron Courville, Yoshua Bengio Generative Adversarial Nets. Advances in Neural Information Processing Systems: NeurIPS Proceedings 2014.
24 Diederik P Kingma MW. Auto-Encoding Variational Bayes. *ICLR*. 2014.
25 Baur C, Wiestler B, Albarqouni S, Navab N. Scale-Space Autoencoders for Unsupervised Anomaly Segmentation in Brain MRI. Cham: Springer International Publishing; 2020:552-561.
26 Jiang J. Robust prediction of immune checkpoint inhibition therapy for non-small cell lung cancer. Frontiers in Immunology 2021; 12: 646874.
27 Barber PR, Mustapha R, Flores-Borja F et al. Predicting progression-free survival after systemic therapy in advanced head and neck cancer: Bayesian regression and model development. eLife 2022; 11: e73288.
28 Barber PR, Weitsman G, Lawler K et al. HER2-HER3 Heterodimer Quantification by FRET-FLIM and Patient Subclass Analysis of the COIN Colorectal Trial. Journal of the National Cancer Institute 2020; 112: 944-954.
